# Microbiome Profiling Reveals Prognostic Heterogeneity in *Staphylococcus aureus* Pneumonia

**DOI:** 10.64898/2026.09.17.26363347

**Authors:** Georgios D. Kitsios, Michael Aaron Sy, William G. Bain, Matthew Hensley, Shulin Qin, Xiaohong Wang, Kyle Inman, Charles Dela Cruz, Keven Robinson, Seyed Mehdi Nouraie, Faraaz A. Shah, Panayiotis V. Benos, Bryan J. McVerry, Alison Morris

**Author notes:** Corresponding author: Georgios D. Kitsios, MD, PhD Assistant Professor of Medicine, Division of Pulmonary, Allergy, Critical Care and Sleep Medicine University of Pittsburgh Medical Center, Address: UPMC Montefiore Hospital, NW628, 3459 Fifth Avenue, Pittsburgh, PA 15213. Co-senior authors (Bryan J. McVerry, Alison Morris).

## Abstract

**Background:** Staphylococcus aureus is a leading cause of severe pneumonia in mechanically ventilated patients. Clinical cultures identify pathogen presence but may not reflect lower respiratory tract microbial ecology. Whether culture-confirmed S. aureus pneumonia encompasses compositional heterogeneity with prognostic implications remains unknown.

**Methods:** We performed 16S rRNA gene sequencing and shotgun nanopore metagenomics on endotracheal aspirate samples from mechanically ventilated patients with culture-confirmed S. aureus pneumonia in a prospective ICU registry. We quantified Staphylococcus abundance, assessed correlations with culture characteristics and host inflammatory biomarkers, and examined associations with 60-day mortality using Kaplan-Meier and Cox hazards analyses.

**Results:** Among 109 patients, semi-quantitative culture growth and methicillin resistance showed no associations with outcomes. 16S sequencing (n=54) revealed marked heterogeneity in Staphylococcus relative abundance (range 0–96.7%), with only 33% demonstrating dominance (>50%). Dominance was associated with worse 60-day survival (50% vs. 80%,p=0.013) and remained independently predictive after adjusting for age, sex, and methicillin resistance (adjusted HR 3.24 [95%CI 1.12–9.36],p=0.030). Patients with dominance exhibited elevated pentraxin-3 (p=0.01) and reduced fractalkine (p=0.02). Nanopore metagenomics (n=28) validated these findings, with high absolute S. aureus read counts independently predicting mortality (adjusted HR 11.23 [95%CI 2.25–55.9],p=0.003). In an exploratory analysis of virulence genes (n=19), staphylokinase detection was associated with the hyperinflammatory phenotype (p=0.003) and mortality (p=0.046).

**Conclusions:** Metagenomic profiling reveals clinically meaningful heterogeneity within culture-confirmed S. aureus pneumonia, masked by conventional diagnostics. Staphylococcus dominance identifies a high-risk phenotype with elevated bacterial burden, dysregulated host responses, and increased mortality, challenging the assumption that culture positivity represents a uniform clinical entity.

## Introduction

Pneumonia remains a leading cause of sepsis, acute respiratory distress syndrome, and mortality in critically ill patients requiring mechanical ventilation.(1, 2) Despite advances in critical care, timely and accurate identification of causative pathogens continues to challenge clinicians, as current diagnostic approaches rely heavily on microbiological culture—a technology that, while considered the reference standard, suffers from well-documented limitations including slow turnaround times, low sensitivity, and susceptibility to overgrowth bias.(3–5) These constraints directly impact antimicrobial stewardship and patient outcomes, as treatment decisions must often be made empirically while awaiting culture results that may ultimately prove uninformative.

Metagenomic next-generation sequencing (mNGS) offers the theoretical advantages of rapid, comprehensive, and hypothesis-free pathogen detection directly from respiratory specimens. Clinical studies have demonstrated that metagenomics can identify pathogens missed by conventional cultures, detect polymicrobial infections, and characterize antimicrobial resistance genes.(6–8) However, routine clinical adoption remains constrained by lack of standardized analytical pipelines and interpretation criteria, persistent difficulty distinguishing colonization from infection in non-sterile respiratory samples, and limited prospective evidence that improved pathogen detection translates into better patient-centered outcomes.(9)

A fundamental challenge in validating mNGS for pneumonia diagnosis is that performance metrics are necessarily benchmarked against culture—the very imperfect reference standard mNGS aims to improve.(5) This circular logic assumes that culture-positive cases represent uniform “true positives” against which to calibrate abundance thresholds and interpretation criteria for culture-negative cases.(10) However, before addressing culture’s false-negative rate, a more fundamental question merits examination: among culture-positive cases presumed to represent diagnostic certainty, does the reference standard itself accurately reflect the underlying microbial ecology? *Staphylococcus aureus* (*S. aureus*) pneumonia provides an instructive model to interrogate this assumption.

As one of the most common and lethal pathogens in severe community- or hospital-acquired pneumonia, *S. aureus* mandates pathogen-specific antimicrobial therapy distinct from empiric regimens.(11) The clinical observation that *S. aureus* demonstrates robust growth on routine culture media(12)—captured in the dictum that “Staph is not shy”—suggests the organism’s dominance in clinical specimens.(13, 14) However, whether vigorous culture growth accurately reflects the true microbial ecology of the lower respiratory tract (LRT), or represents selective amplification of a single organism during culture, remains unexplored. If culture-positive *S. aureus* pneumonia encompasses substantial ecological heterogeneity masked by culture techniques, this variation may have important implications for disease pathogenesis, clinical outcomes, and therapeutic strategies.

In this study, we used culture-confirmed *S. aureus* pneumonia as a defined clinical entity to systematically compare conventional microbiologic methods with metagenomic approaches. We hypothesized that mNGS profiling would uncover compositional heterogeneity with prognostic relevance even within patients meeting current gold-standard diagnostic criteria for S. aureus pneumonia. Using 16S rRNA gene amplicon sequencing and Nanopore mNGS from endotracheal aspirates (ETAs) of mechanically ventilated patients with culture-confirmed *S. aureus* pneumonia, we characterized microbiota profiles, assessed their relationship to host inflammatory responses and clinical outcomes, and evaluated the potential of mNGS approaches for biological phenotyping beyond conventional culture results.

## Methods

### Study Design and Population

This was a nested cohort study conducted within a prospective registry of critically ill patients with acute respiratory failure requiring invasive mechanical ventilation. The parent cohort is the Acute Lung Injury Registry and Biospecimen Repository (ALIR), which enrolls patients from UPMC Hospitals ICUs. The study was approved by the University of Pittsburgh Institutional Review Board (STUDY19050099), and informed consent was obtained from patients or their legally authorized representatives. All procedures followed the ethical standards of the Declaration of Helsinki. For this nested study, we identified adult patients (18-90 years) admitted to medical or cardiac intensive care units who met the following inclusion criteria: (1) endotracheal intubation, and (2) positive LRT culture for S. aureus from ETA or bronchoalveolar lavage fluid (BALF) obtained within 72 hours before or after intubation, with any amount of growth reported by clinical microbiology. This 72-hour time window was chosen to capture patients with community- or hospital-acquired pneumonia severe enough to require mechanical ventilation, while excluding ventilator-associated pneumonia (VAP) that develops after prolonged intubation. We excluded patients with chronic ventilator dependence via tracheostomy. Of 109 patients meeting these criteria, subsets underwent 16S rRNA gene (n=54) and Nanopore sequencing (n=28); these subsets reflect random selection based on biospecimen and sequencing capacity availability rather than differential clinical eligibility and were drawn from the same parent cohort of 109 culture-confirmed cases.

### Clinical Microbiology Testing

LRT specimens (ETA or BALF, obtained per standard clinical care rather than a study-specific research protocol) were processed by the UPMC clinical microbiology laboratory using standard protocols. ETA specimens underwent semi-quantitative culture with growth reported as rare, light, moderate, or heavy. BALF specimens obtained via bronchoscopy were cultured using either quantitative or semi-quantitative methods. Quantitative results were converted to semi-quantitative categories to standardize growth assessments across all specimens. *S. aureus* isolates were classified as methicillin-susceptible (MSSA) or methicillin-resistant (MRSA) using standard susceptibility testing methods. We recorded the presence and semi-quantitative amount of *S. aureus* growth and documented all co-detected organisms, including oral commensals (often reported as normal respiratory flora), yeast, and other respiratory bacterial or fungal pathogens. For analysis, culture patterns were categorized into three clinically relevant groups: (1) *S. aureus* only, (2) *S. aureus* with other bacterial pathogens (gram-negative or gram-positive organisms, or polymicrobial infections), and (3) *S. aureus* with organisms not considered typical respiratory pathogens (i.e. normal respiratory flora or yeast). These categories have direct implications for antimicrobial therapy, with groups 1 and 3 requiring anti-staphylococcal coverage alone, while group 2 requires broader-spectrum therapy.

### Biospecimen Collection and Processing

Research biospecimens were collected within 72 hours of intubation. ETAs were obtained by suctioning secretions from the lower airway via the endotracheal tube using an in-line suction catheter without breaking the ventilatory circuit seal. Oral swabs were obtained by gently swabbing the posterior oropharynx with a sterile cotton swab for 5 seconds. Simultaneous blood samples were collected for plasma separation.

### 16S rRNA Gene Sequencing

In a random subset of 54 participants with available ETA biospecimens, we extracted genomic DNA and performed 16S rRNA gene amplicon sequencing of the V4 hypervariable region on the Illumina MiSeq platform. Oral swabs collected at the same baseline timepoint were processed and sequenced using the same protocol to enable paired oral-lower respiratory tract comparisons in a subset of patients. Experimental negative controls and mock microbial community positive controls (Zymo Research) were included in all processing steps. Sequences were processed with a Mothur-based pipeline as previously described.(15) Taxonomic classifications were performed using the Ribosomal Database Project’s naïve Bayesian classifier with the SILVA 16S rRNA database (v138).(15, 16)

### Nanopore Metagenomic Sequencing

In a subset of 28 participants, we performed shotgun mNGS of ETA samples using Oxford Nanopore Technologies. Host DNA depletion was performed using a saponin-based method. Libraries of 6-12 samples per run were prepared using a rapid PCR barcoding kit (SQK-RPB004 or SQK-RPB114.24) and sequenced on MinION devices (Mk1b and Mk1c flowcells) for up to 24 hours.(15, 16) FASTQ files were quality filtered (mean q-score ≥7) and processed through the BugSeq bioinformatics platform for taxonomic classification, antimicrobial resistance gene detection, and virulence factor identification.(17)

Virulence gene content was characterized from Nanopore data using BugSeq taxonomic and functional annotation. Among participants with sufficient sequencing depth for functional annotation (n=19), virulence gene profiles were catalogued at the individual sample level; detected gene categories included delta-hemolysin (*hld*), staphylococcal enterotoxins, and staphylokinase (*sak*). Based on the biological rationale that staphylokinase detection marks a distinct high-virulence clonal lineage — given its known role in immune evasion via IgG and complement cleavage and its enrichment in community-acquired *S. aureus* lineages — participants were dichotomized into staphylokinase-positive (n=6) versus staphylokinase-negative (n=13) groups. All virulence gene classifications were derived from metagenomic reads and represent gene presence/detection rather than confirmed protein expression.

### Host Inflammatory Biomarker Measurements

We measured inflammatory and lung injury biomarkers in ETA supernatants and plasma samples using a custom 10-plex Luminex panel (R&D Systems). Biomarkers included interleukin (IL)-6, IL-8, IL-10, soluble tumor necrosis factor receptor 1 (sTNFR1), suppressor of tumorigenicity-2 (ST2), fractalkine, soluble receptor for advanced glycation end-products (sRAGE), angiopoietin-2, procalcitonin, and pentraxin-3.(15, 16)

### Statistical Analyses

We performed non-parametric comparisons for continuous variables (described as median and interquartile range) and categorical variables using Wilcoxon rank-sum and Fisher’s exact tests, respectively, including for associations between staphylokinase status and airway and systemic inflammatory biomarkers or clinical outcomes, given non-normal distributions and small subgroup sizes. From plasma biomarkers, we classified participants into a hyperinflammatory vs. hypoinflammatory phenotype using predictions from a logistic regression model utilizing procalcitonin, Ang-2, sTNFR1 and bicarbonate levels.(18) For microbiome analyses, we included samples after filtering for singletons and low-abundance taxa (relative abundance <0.0001 in <5% of samples). We analyzed 16S rRNA gene sequences at the genus level and Nanopore metagenomic sequences at the species level. Alpha diversity was calculated using the Shannon index and Beta diversity was assessed using Euclidean distances on centered log-ratio [CLR] transformed abundances, analyzed via permutation analysis of variance (PERMANOVA), and visualized with principal coordinates analysis (PCoA). We defined *Staphylococcus* dominance as relative abundance >50% in 16S profiles consistent with thresholds used in related pneumonia cohorts(19, 20), and high *S. aureus* abundance as upper tertile (>2,423 reads) in Nanopore profiles. Survival was assessed at 60 days post-enrollment using Kaplan-Meier curves compared with log-rank tests and Cox proportional hazards models adjusting for age, sex, and MRSA status. We examined associations between microbiota profiles and host inflammatory biomarkers using Spearman correlations. All statistical tests were two-sided with significance threshold of α=0.05. Analyses were performed using R version 4.4.0 with vegan, survival, and mia packages.

## Results

### Clinical Microbiologic Testing

Between October 2011 and October 2023, we enrolled 109 participants with culture-confirmed *S. aureus* pneumonia requiring mechanical ventilation. All patients received antibiotics during the study period, with 97 (89.0%) receiving anti-staphylococcal coverage and 96 (88.1%) receiving anti-MRSA antibiotics.

Culture growth patterns demonstrated variable *S. aureus* burden, with rare growth in 7 (6.4%), light in 25 (22.9%), moderate in 33 (30.3%), and heavy growth in 44 (40.4%) cases. Co-infection patterns revealed that *S. aureus* was isolated alone in 24 (22.0%) cases, with other bacterial pathogens in 38 (34.9%) cases, and with non-pathogenic organisms (normal respiratory flora or yeast) in 47 (43.1%) cases (Table 1). Forty-six (42.2%) cases were diagnosed with MRSA based on antibiograms and the remainder with MSSA (Table S1). Patients with *S. aureus* isolated alone were significantly more likely to have concurrent bacteremia compared to those with co-detected organisms (33.3% vs. 13.2% vs. 4.3%, p<0.01). There were no significant differences in host inflammatory biomarkers, ventilator parameters or outcomes across co-infection pattern groups (Table 1 and Figure S1).

**Table 1.**
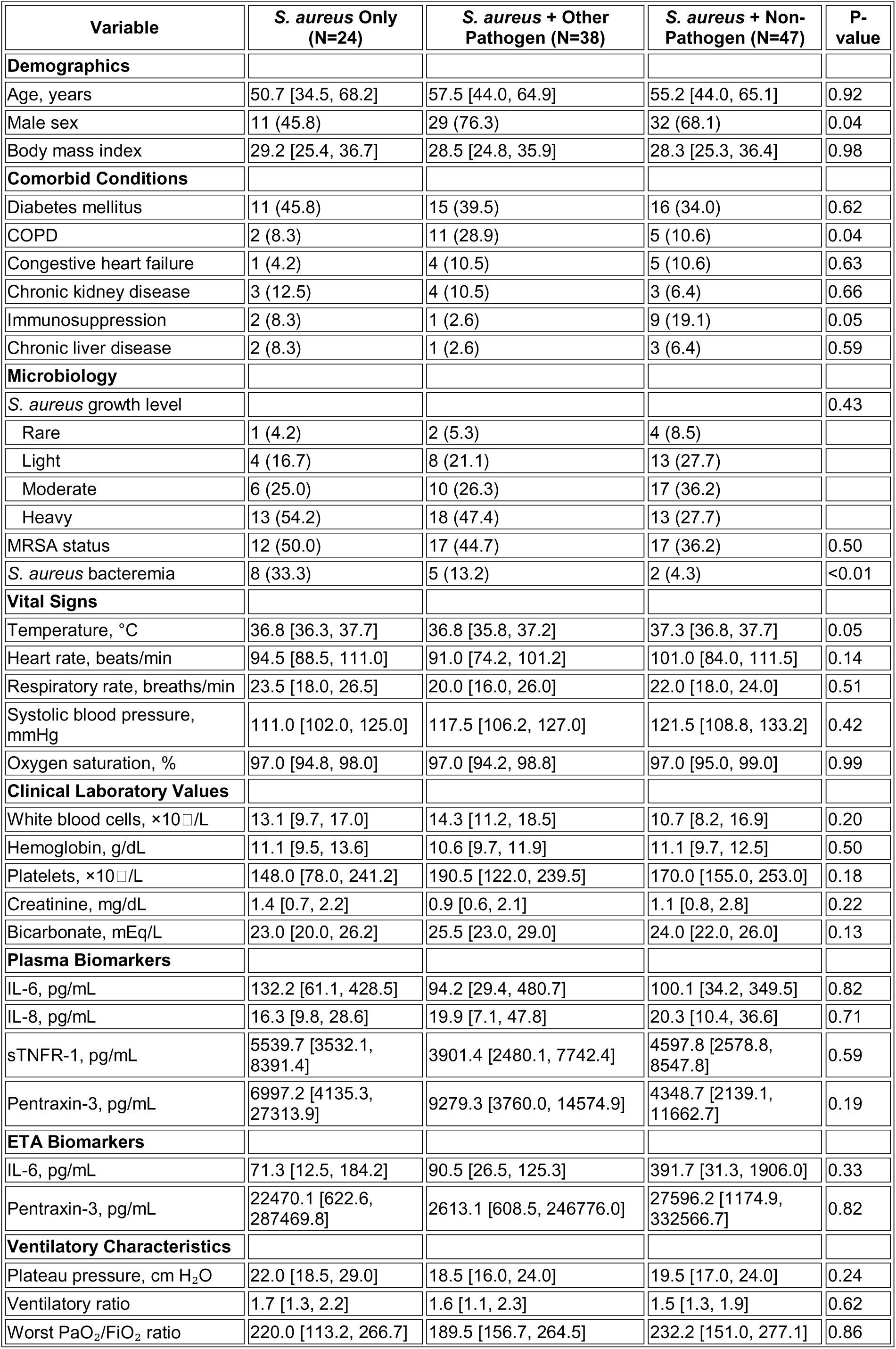
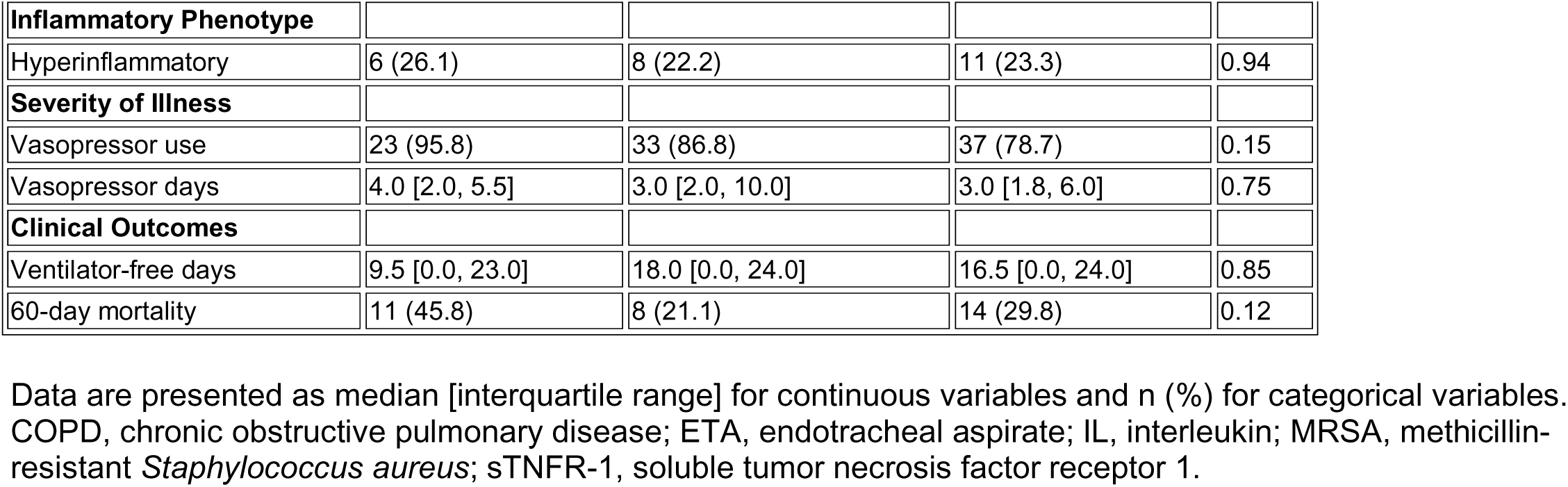
Baseline Characteristics Stratified by Co-Infection Pattern.

| Variable | *S. aureus* Only (N=24) | *S. aureus* + Other Pathogen (N=38) | *S. aureus* + Non-Pathogen (N=47) | P-value |
| --- | --- | --- | --- | --- |
| **Demographics** |  |  |  |  |
| Age, years | 50.7 [34.5, 68.2] | 57.5 [44.0, 64.9] | 55.2 [44.0, 65.1] | 0.92 |
| Male sex | 11 (45.8) | 29 (76.3) | 32 (68.1) | 0.04 |
| Body mass index | 29.2 [25.4, 36.7] | 28.5 [24.8, 35.9] | 28.3 [25.3, 36.4] | 0.98 |
| **Comorbid Conditions** |  |  |  |  |
| Diabetes mellitus | 11 (45.8) | 15 (39.5) | 16 (34.0) | 0.62 |
| COPD | 2 (8.3) | 11 (28.9) | 5 (10.6) | 0.04 |
| Congestive heart failure | 1 (4.2) | 4 (10.5) | 5 (10.6) | 0.63 |
| Chronic kidney disease | 3 (12.5) | 4 (10.5) | 3 (6.4) | 0.66 |
| Immunosuppression | 2 (8.3) | 1 (2.6) | 9 (19.1) | 0.05 |
| Chronic liver disease | 2 (8.3) | 1 (2.6) | 3 (6.4) | 0.59 |
| **Microbiology** |  |  |  |  |
| *S. aureus* growth level |  |  |  | 0.43 |
| Rare | 1 (4.2) | 2 (5.3) | 4 (8.5) |  |
| Light | 4 (16.7) | 8 (21.1) | 13 (27.7) |  |
| Moderate | 6 (25.0) | 10 (26.3) | 17 (36.2) |  |
| Heavy | 13 (54.2) | 18 (47.4) | 13 (27.7) |  |
| MRSA status | 12 (50.0) | 17 (44.7) | 17 (36.2) | 0.50 |
| *S. aureus* bacteremia | 8 (33.3) | 5 (13.2) | 2 (4.3) | <0.01 |
| **Vital Signs** |  |  |  |  |
| Temperature, °C | 36.8 [36.3, 37.7] | 36.8 [35.8, 37.2] | 37.3 [36.8, 37.7] | 0.05 |
| Heart rate, beats/min | 94.5 [88.5, 111.0] | 91.0 [74.2, 101.2] | 101.0 [84.0, 111.5] | 0.14 |
| Respiratory rate, breaths/min | 23.5 [18.0, 26.5] | 20.0 [16.0, 26.0] | 22.0 [18.0, 24.0] | 0.51 |
| Systolic blood pressure, mmHg | 111.0 [102.0, 125.0] | 117.5 [106.2, 127.0] | 121.5 [108.8, 133.2] | 0.42 |
| Oxygen saturation, % | 97.0 [94.8, 98.0] | 97.0 [94.2, 98.8] | 97.0 [95.0, 99.0] | 0.99 |
| **Clinical Laboratory Values** |  |  |  |  |
| White blood cells, ×10<sup>3</sup>/L | 13.1 [9.7, 17.0] | 14.3 [11.2, 18.5] | 10.7 [8.2, 16.9] | 0.20 |
| Hemoglobin, g/dL | 11.1 [9.5, 13.6] | 10.6 [9.7, 11.9] | 11.1 [9.7, 12.5] | 0.50 |
| Platelets, ×10<sup>3</sup>/L | 148.0 [78.0, 241.2] | 190.5 [122.0, 239.5] | 170.0 [155.0, 253.0] | 0.18 |
| Creatinine, mg/dL | 1.4 [0.7, 2.2] | 0.9 [0.6, 2.1] | 1.1 [0.8, 2.8] | 0.22 |
| Bicarbonate, mEq/L | 23.0 [20.0, 26.2] | 25.5 [23.0, 29.0] | 24.0 [22.0, 26.0] | 0.13 |
| **Plasma Biomarkers** |  |  |  |  |
| IL-6, pg/mL | 132.2 [61.1, 428.5] | 94.2 [29.4, 480.7] | 100.1 [34.2, 349.5] | 0.82 |
| IL-8, pg/mL | 16.3 [9.8, 28.6] | 19.9 [7.1, 47.8] | 20.3 [10.4, 36.6] | 0.71 |
| sTNFR-1, pg/mL | 5539.7 [3532.1, 8391.4] | 3901.4 [2480.1, 7742.4] | 4597.8 [2578.8, 8547.8] | 0.59 |
| Pentraxin-3, pg/mL | 6997.2 [4135.3, 27313.9] | 9279.3 [3760.0, 14574.9] | 4348.7 [2139.1, 11662.7] | 0.19 |
| **ETA Biomarkers** |  |  |  |  |
| IL-6, pg/mL | 71.3 [12.5, 184.2] | 90.5 [26.5, 125.3] | 391.7 [31.3, 1906.0] | 0.33 |
| Pentraxin-3, pg/mL | 22470.1 [622.6, 287469.8] | 2613.1 [608.5, 246776.0] | 27596.2 [1174.9, 332566.7] | 0.82 |
| **Ventilatory Characteristics** |  |  |  |  |
| Plateau pressure, cm H<sub>2</sub>O | 22.0 [18.5, 29.0] | 18.5 [16.0, 24.0] | 19.5 [17.0, 24.0] | 0.24 |
| Ventilatory ratio | 1.7 [1.3, 2.2] | 1.6 [1.1, 2.3] | 1.5 [1.3, 1.9] | 0.62 |
| Worst PaO<sub>2</sub>/FiO<sub>2</sub> ratio | 220.0 [113.2, 266.7] | 189.5 [156.7, 264.5] | 232.2 [151.0, 277.1] | 0.86 |

| Variable | *S. aureus* Only<br>(N=24) | *S. aureus* + Other<br>Pathogen (N=38) | *S. aureus* + Non-Pathogen (N=47) | P-value |
| --- | --- | --- | --- | --- |
| **Inflammatory Phenotype** |  |  |  |  |
| Hyperinflammatory | 6 (26.1) | 8 (22.2) | 11 (23.3) | 0.94 |
| **Severity of Illness** |  |  |  |  |
| Vasopressor use | 23 (95.8) | 33 (86.8) | 37 (78.7) | 0.15 |
| Vasopressor days | 4.0 [2.0, 5.5] | 3.0 [2.0, 10.0] | 3.0 [1.8, 6.0] | 0.75 |
| **Clinical Outcomes** |  |  |  |  |
| Ventilator-free days | 9.5 [0.0, 23.0] | 18.0 [0.0, 24.0] | 16.5 [0.0, 24.0] | 0.85 |
| 60-day mortality | 11 (45.8) | 8 (21.1) | 14 (29.8) | 0.12 |

Data are presented as median [interquartile range] for continuous variables and n (%) for categorical variables. COPD, chronic obstructive pulmonary disease; ETA, endotracheal aspirate; IL, interleukin; MRSA, methicillin-resistant *Staphylococcus aureus*; sTNFR-1, soluble tumor necrosis factor receptor 1.

### Bacterial profiling by 16S sequencing

We performed 16S rRNA gene sequencing on 54 available ETA samples. *Staphylococcus* genus relative abundance demonstrated striking heterogeneity, ranging from 0% to 96.7% (Figure 1A), despite all cases meeting standard microbiological criteria for *S. aureus* pneumonia. Eighteen (33.3%) cases exceeded the operationally-defined threshold (>50% relative abundance) for *Staphylococcus* dominance. Comparison of patients with *Staphylococcus* dominance versus those without revealed no significant differences in demographics, comorbidities, severity of illness, ventilator parameters, or most laboratory values (Table 2), though BUN was significantly higher in dominant cases (43.5 vs. 27.5 mg/dL, p=0.02) and ICU mortality was significantly higher (38.9% vs. 8.3%, p=0.02).

**Figure 1.**
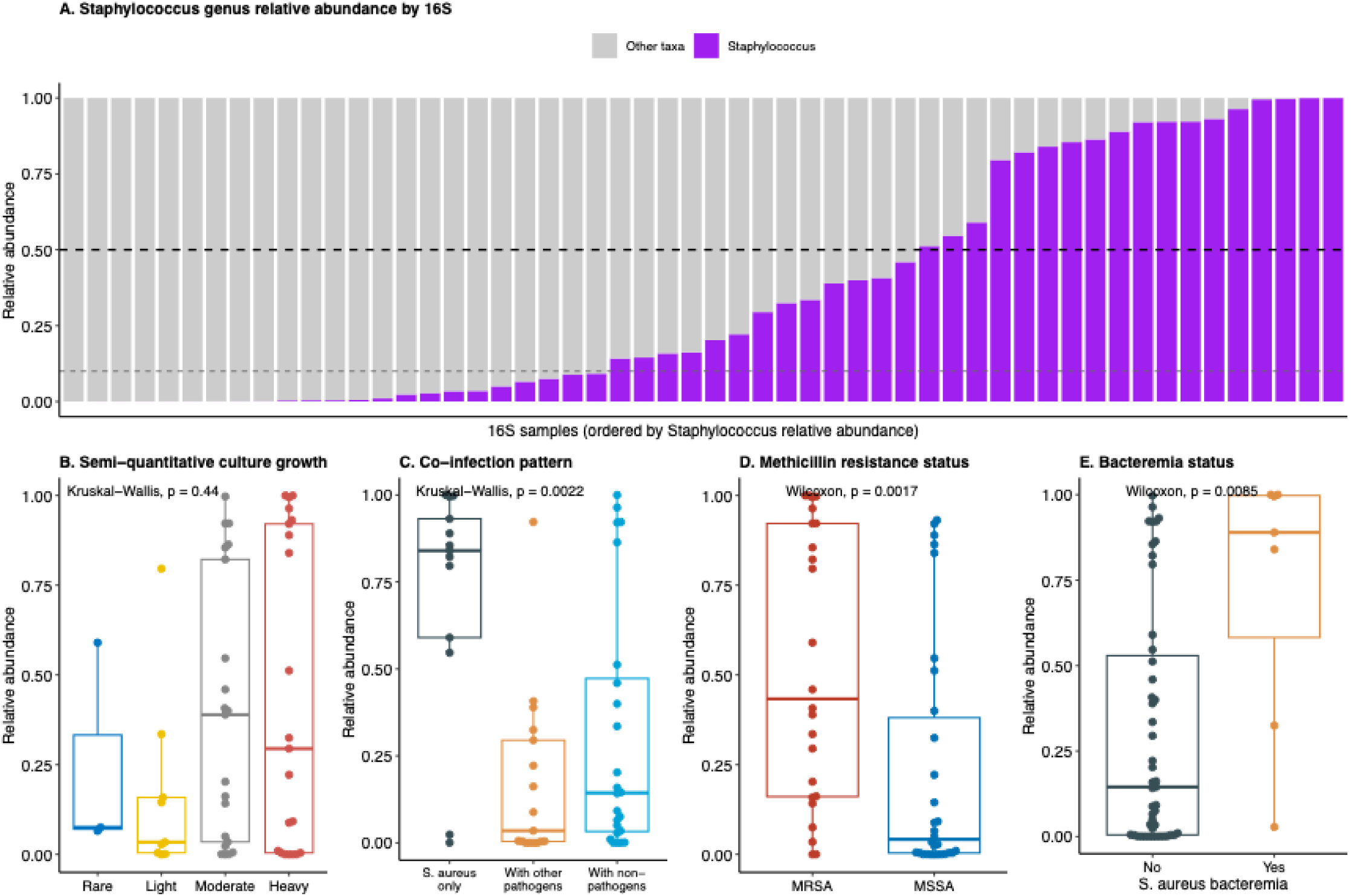
Staphylococcus Relative Abundance by 16S rRNA Gene Sequencing Shows Marked Heterogeneity and Associates with Clinical Culture Characteristics. (A) Stacked bar plot showing *Staphylococcus* genus relative abundance (purple) and other taxa (gray) in endotracheal aspirate samples from 54 patients with culture-confirmed *S. aureus* pneumonia, ordered by relative abundance. Thick dashed line indicates the 50% dominance threshold; thin dashed line indicates 10% relative abundance. (B) Relative abundance by semi-quantitative culture growth level, showing no significant association (Kruskal-Wallis p=0.44). (C) Relative abundance by co-infection pattern, showing significantly higher abundance in *S. aureus*-only cases (Kruskal-Wallis p=0.0022). (D) Relative abundance by methicillin resistance status, with MRSA cases showing higher abundance than MSSA (Wilcoxon p=0.0085). (E) Relative abundance in patients with and without *S. aureus* bacteremia (Wilcoxon p=0.0085). Box plots show median, interquartile range, and range with individual points overlaid.

**Table 2.** Baseline Characteristics Stratified by Staphylococcus Dominance in 16S Microbiome Profiles.

| Variable | No Dominance (N=36) | *Staphylococcus* Dominance (N=18) | P-value |
| --- | --- | --- | --- |
| **Demographics** |  |  |  |
| Age, years | 58.4 [44.9, 67.1] | 59.2 [35.7, 70.4] | 0.81 |
| Male sex | 25 (69.4) | 8 (44.4) | 0.14 |
| Body mass index | 28.6 [25.6, 36.4] | 26.1 [24.2, 33.7] | 0.20 |
| **Comorbid Conditions** |  |  |  |
| Diabetes mellitus | 12 (33.3) | 9 (50.0) | 0.37 |
| COPD | 7 (19.4) | 2 (11.1) | 0.70 |
| Congestive heart failure | 3 (8.3) | 2 (11.1) | 1.00 |
| Chronic kidney disease | 4 (11.1) | 3 (16.7) | 0.89 |
| Immunosuppression | 5 (13.9) | 4 (22.2) | 0.70 |
| Chronic liver disease | 3 (8.3) | 2 (11.1) | 1.00 |
| **Microbiology** |  |  |  |
| *S. aureus* growth level |  |  | 0.41 |
| Rare | 2 (5.6) | 1 (5.6) |  |
| Light | 8 (22.2) | 1 (5.6) |  |
| Moderate | 14 (38.9) | 7 (38.9) |  |
| Heavy | 12 (33.3) | 9 (50.0) |  |
| MRSA status | 13 (36.1) | 11 (61.1) | 0.15 |
| *S. aureus* bacteremia | 2 (5.6) | 5 (27.8) | 0.06 |
| **Vital Signs** |  |  |  |
| Temperature, °C | 37.0 [36.3, 37.6] | 37.0 [36.4, 37.3] | 0.80 |
| Heart rate, beats/min | 93.0 [74.8, 103.5] | 101.5 [87.0, 111.0] | 0.19 |
| Respiratory rate, breaths/min | 20.0 [16.8, 25.2] | 20.5 [18.0, 24.0] | 0.79 |
| Systolic blood pressure, mmHg | 123.0 [108.5, 132.0] | 108.0 [95.0, 133.2] | 0.32 |
| Oxygen saturation, % | 97.0 [95.0, 99.0] | 97.0 [97.0, 98.8] | 0.79 |
| **Clinical Laboratory Values** |  |  |  |
| White blood cells, ×10<sup>3</sup>/L | 11.2 [9.1, 16.2] | 14.5 [11.2, 20.4] | 0.16 |
| Hemoglobin, g/dL | 10.6 [9.4, 11.6] | 10.4 [9.2, 11.8] | 0.87 |
| Platelets, ×10<sup>3</sup>/L | 181.5 [156.5, 252.0] | 179.0 [137.5, 244.8] | 0.76 |
| Creatinine, mg/dL | 1.1 [0.8, 1.9] | 1.8 [1.0, 3.4] | 0.07 |
| BUN, mg/dL | 27.5 [18.8, 41.5] | 43.5 [28.0, 71.5] | 0.02 |
| Bicarbonate, mEq/L | 23.5 [22.0, 26.2] | 22.5 [18.2, 24.0] | 0.06 |
| **Plasma Biomarkers** |  |  |  |
| IL-6, pg/mL | 85.0 [20.5, 423.0] | 112.4 [60.4, 157.3] | 0.88 |
| sTNFR-1, pg/mL | 4539.0 [2578.8, 7858.2] | 6471.9 [4016.2, 10072.9] | 0.05 |
| Pentraxin-3, pg/mL | 5768.1 [2576.6, 13353.6] | 8227.0 [4005.7, 14788.3] | 0.39 |
| **ETA Biomarkers** |  |  |  |
| IL-6, pg/mL | 118.0 [54.9, 513.8] | 521.5 [434.7, 4055.9] | 0.15 |
| Pentraxin-3, pg/mL | 36562.2 [1453.3, 162439.3] | 766363.1 [637888.2, 1058801.0] | 0.01 |
| Fractalkine, pg/mL | 1969.6 [26.9, 5157.8] | 26.9 [26.9, 26.9] | 0.02 |
| **Ventilatory Characteristics** |  |  |  |
| Plateau pressure, cm H<sub>2</sub>O | 19.0 [15.5, 22.0] | 16.0 [14.0, 21.0] | 0.46 |
| Ventilatory ratio | 1.6 [1.4, 1.9] | 1.5 [1.3, 1.8] | 0.93 |
| Worst PaO<sub>2</sub>/FiO<sub>2</sub> ratio | 193.7 [151.9, 280.8] | 247.5 [223.1, 263.9] | 0.54 |
| **Inflammatory Phenotype** |  |  |  |
| Hyperinflammatory | 8 (22.2) | 7 (38.9) | 0.25 |
| Severity of Illness |  |  |  |
| Vasopressor use | 27 (75.0) | 17 (94.4) | 0.17 |
| Vasopressor days | 4.0 [2.0, 5.5] | 6.0 [1.0, 6.0] | 0.68 |
| Clinical Outcomes |  |  |  |
| Ventilator-free days | 19.5 [11.8, 24.0] | 9.5 [0.0, 23.2] | 0.16 |
| ICU mortality | 3 (8.3) | 7 (38.9) | 0.02 |
| 60-day mortality | 7 (19.4) | 9 (50.0) | 0.05 |

Data are presented as median [interquartile range] for continuous variables and n (%) for categorical variables. BUN, blood urea nitrogen; COPD, chronic obstructive pulmonary disease; ETA, endotracheal aspirate; IL, interleukin; MRSA, methicillin-resistant *Staphylococcus aureus*; sTNFR-1, soluble tumor necrosis factor receptor 1.

In comparisons of 16S profiles with clinical culture results, semi-quantitative culture growth levels showed no association with *Staphylococcus* relative abundance (Figure 1B, p=0.44). Culture co-infection patterns demonstrated significant associations with microbiota composition (Figure 1C, p=0.002), with *S. aureus* isolated alone showing higher *Staphylococcus* relative abundance compared to co-detection with other bacterial pathogens or non-pathogenic organisms. MRSA cases demonstrated higher *Staphylococcus* relative abundance compared to MSSA (Figure 1D, p=0.0085). Patients with *S. aureus* bacteremia demonstrated significantly higher *Staphylococcus* relative abundance in ETA compared to those without bacteremia (Figure 1E, p=0.0085).

To assess the biological significance of microbiota composition, we examined systemic and LRT biomarkers in relation to 16S profiles. Patients with hyperinflammatory subphenotype showed numerically higher *Staphylococcus* relative abundance compared to hypoinflammatory patients (Figure 2A, p=0.081). *Staphylococcus* dominance was associated with a trend toward higher plasma sTNFR-1 levels (Figure 2B, p=0.054), significantly elevated ETA pentraxin-3 (Figure 2C, p=9×10⁻), and markedly reduced ETA fractalkine (Figure 2D, p=0.0077).

**Figure 2.**
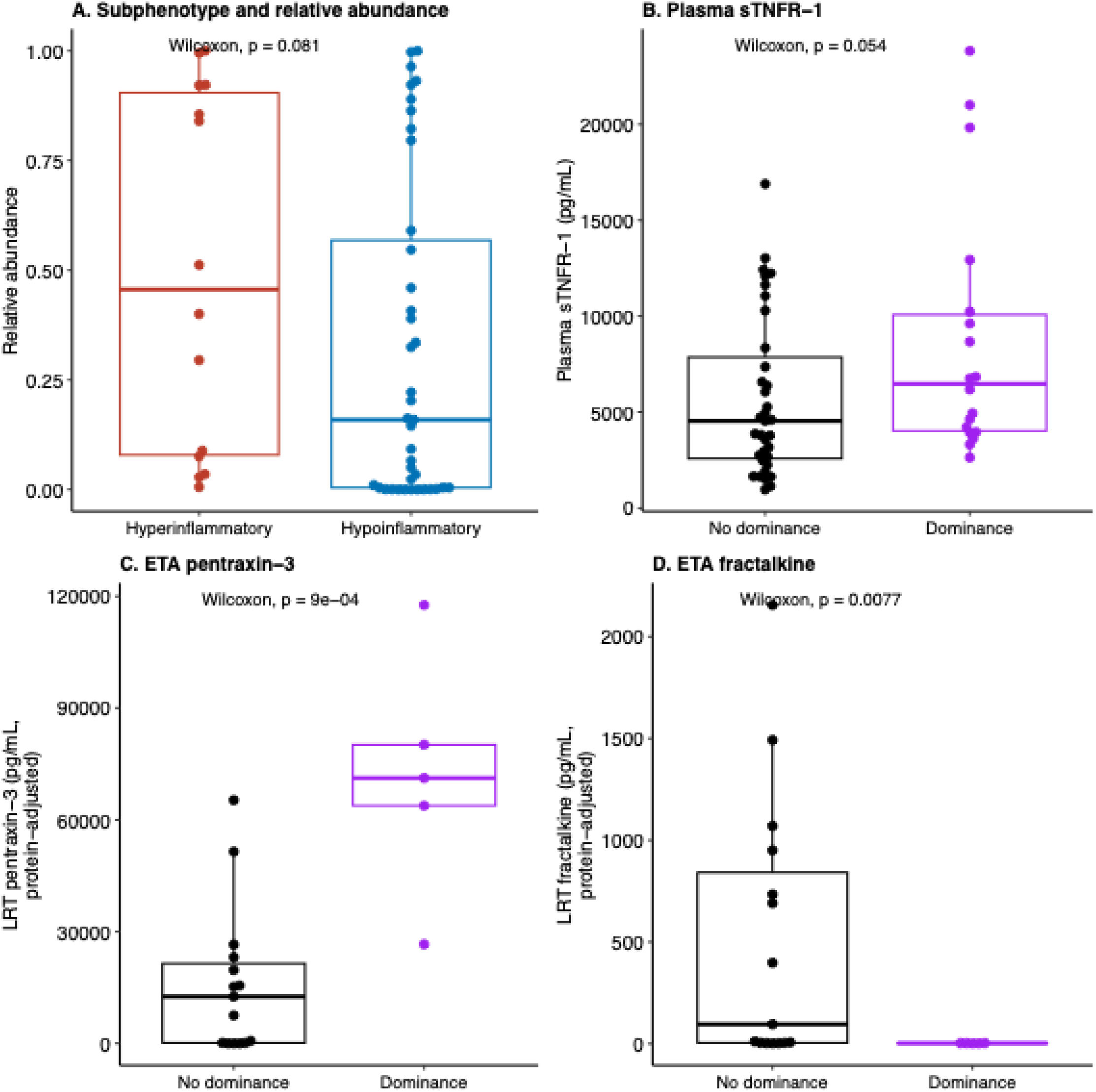
*Staphylococcus* Relative Abundance and Dominance Associate with Host Inflammatory Responses. (A) *Staphylococcus* relative abundance by inflammatory subphenotype (Wilcoxon p=0.081). (B) Plasma sTNFR-1 by dominance status (Wilcoxon p=0.054). (C) ETA pentraxin-3 by dominance status (Wilcoxon p=9×10⁻). (D) ETA fractalkine by dominance status (Wilcoxon p=0.0077). Box plots show median, interquartile range, and range with individual points overlaid.

### Microbiota Composition Associates with Clinical Outcomes

We examined microbiota diversity metrics in relation to 60-day survival. Shannon diversity index showed no significant difference between survivors and non-survivors (Figure 3A, p=0.15). Euclidean distance-based PERMANOVA revealed no significant difference in overall microbial community structure between survivors and non-survivors (p=0.31, Figure 3B). *Staphylococcus* dominance, however, was strongly associated with mortality. Patients with *Staphylococcus* dominance demonstrated 50.0% survival compared to 80.0% in those without dominance (log-rank p=0.013, Figure 3C). In multivariable Cox proportional hazards analysis adjusting for age, sex, and methicillin resistance status, *Staphylococcus* dominance remained independently associated with 60-day mortality (adjusted hazards ratio - HR 3.24 [95% CI 1.12–9.36], p=0.030). In a subset of patients with serial sampling (n=19), non-survivors demonstrated significantly higher baseline *Staphylococcus* relative abundance compared to survivors (p=0.01, Figure S2). This difference was not maintained at the middle timepoint (days 3–6, p=0.11), suggesting that baseline microbiota composition, rather than temporal dynamics, was most strongly associated with outcome.

**Figure 3.**
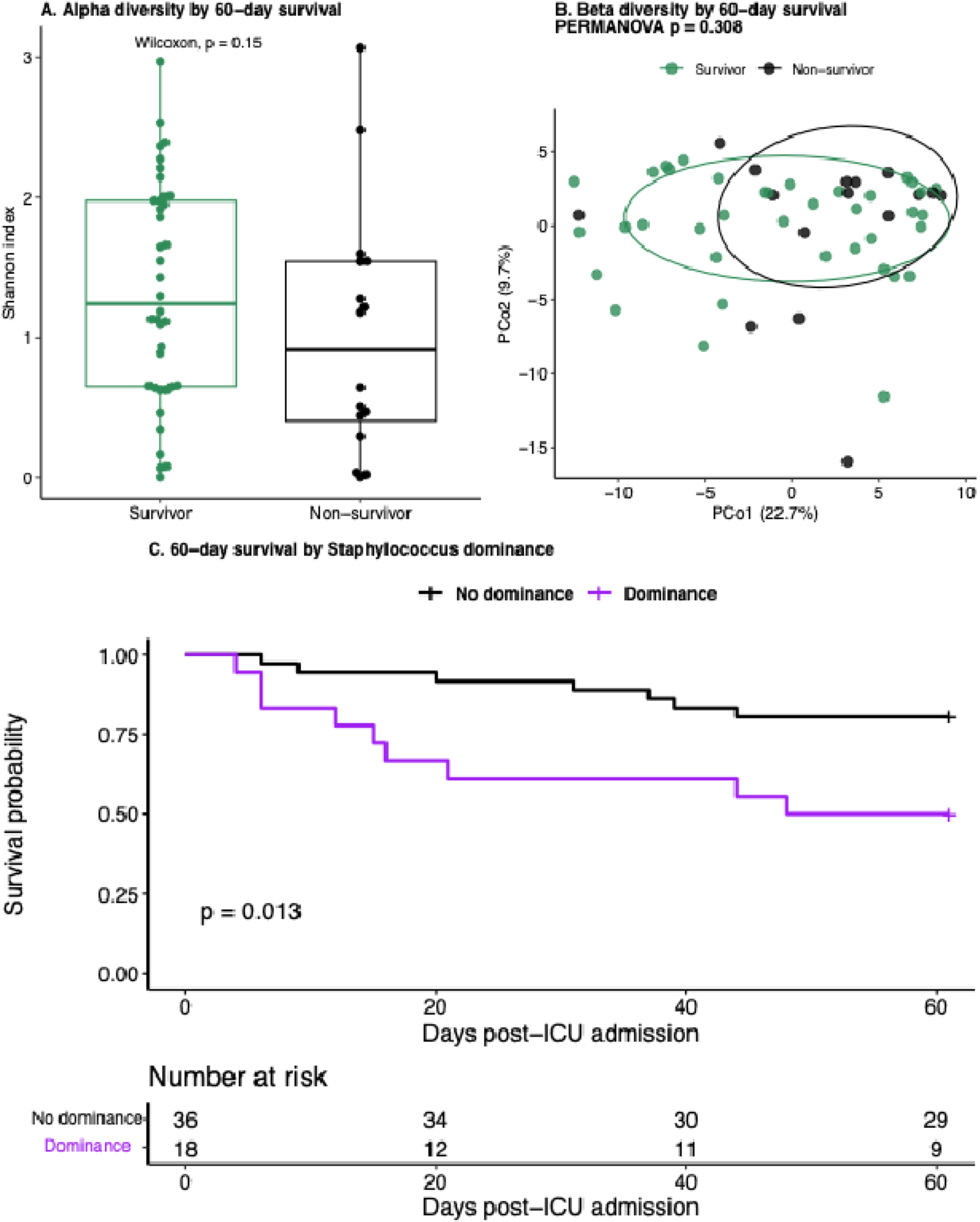
Microbiota Diversity and Staphylococcus Dominance in Relation to 60-Day Survival. (A) Shannon alpha diversity index by 60-day survival status (Wilcoxon p=0.15). (B) Principal coordinates analysis of Aitchison distance (Euclidean distance on centered log-ratio transformed abundances) by 60-day survival status; PERMANOVA revealed no significant difference in community structure (p=0.31). (C) Kaplan-Meier survival curves for 60-day mortality stratified by *Staphylococcus* dominance. Patients with dominance (purple, n=18) demonstrated significantly worse survival than those without (black, n=36), with 50% versus 80% survival at 60 days (log-rank p=0.013). Numbers at risk shown below.

### Oral Microbiome Comparison

To assess whether upper respiratory tract sampling could serve as a non-invasive proxy for LRT *Staphylococcus* burden, we compared paired oral and ETA 16S profiles in 42 patients with baseline samples from both compartments. Oral and ETA *Staphylococcus* relative abundance were modestly but significantly correlated (Spearman ρ=0.361, p=0.019, Figure S3A-B). However, oral *Staphylococcus* dominance (>50%) showed no association with 60-day mortality (log-rank p=0.90, Figure S3C), in contrast to the robust prognostic signal observed for ETA dominance in the same patients (Fisher’s exact OR=4.32, p=0.059). Oral *Staphylococcus* abundance correlated with plasma sTNFR-1 (ρ=0.339, p=0.03) but not with LRT-specific biomarkers (pentraxin-3, fractalkine) or mortality, suggesting oral colonization reflects systemic inflammatory burden without capturing the compartment-specific biology driving clinical outcomes. These findings indicate that oral swabs, while weakly correlated with lung *Staphylococcus* abundance, do not substitute for direct LRT sampling in prognostic assessment.

### Nanopore Metagenomic Sequencing Validates 16S Findings

We performed nanopore mNGS on 28 ETA samples from patients meeting the same baseline culture-confirmation criteria as the parent cohort, to provide orthogonal validation of 16S findings at species-level resolution and to assess antimicrobial resistance genes and virulence factors. Cross-tabulation of *S. aureus* species-level abundance (Nanopore) versus *Staphylococcus* genus-level abundance (16S) demonstrated concordance in samples with both methods available (Figure S4). *S. aureus* absolute abundance demonstrated marked heterogeneity across the cohort, ranging from undetectable to >1.6 million reads (Figure 5A), consistent with 16S findings. In this smaller cohort, semi-quantitative culture growth level, co-infection pattern, and MRSA status showed no significant associations with Nanopore-derived *S. aureus* abundance (all p>0.10, Figure S5B).

We stratified patients by tertiles of absolute *S. aureus* read counts to assess prognostic significance of bacterial burden (low/medium: ≤2,714 reads; high: >2,714 reads). Patients in the high abundance tertile demonstrated significantly worse 60-day survival compared to those in the low/medium tertile (55.6% vs. 89.5% survival, log-rank p=0.008, Figure 4B). In multivariable Cox proportional hazards analysis adjusting for age, sex, and methicillin resistance status, high *S. aureus* abundance remained independently associated with mortality risk (adjusted HR 11.23 [95% CI 2.25–55.9], p=0.003).

**Figure 4.**
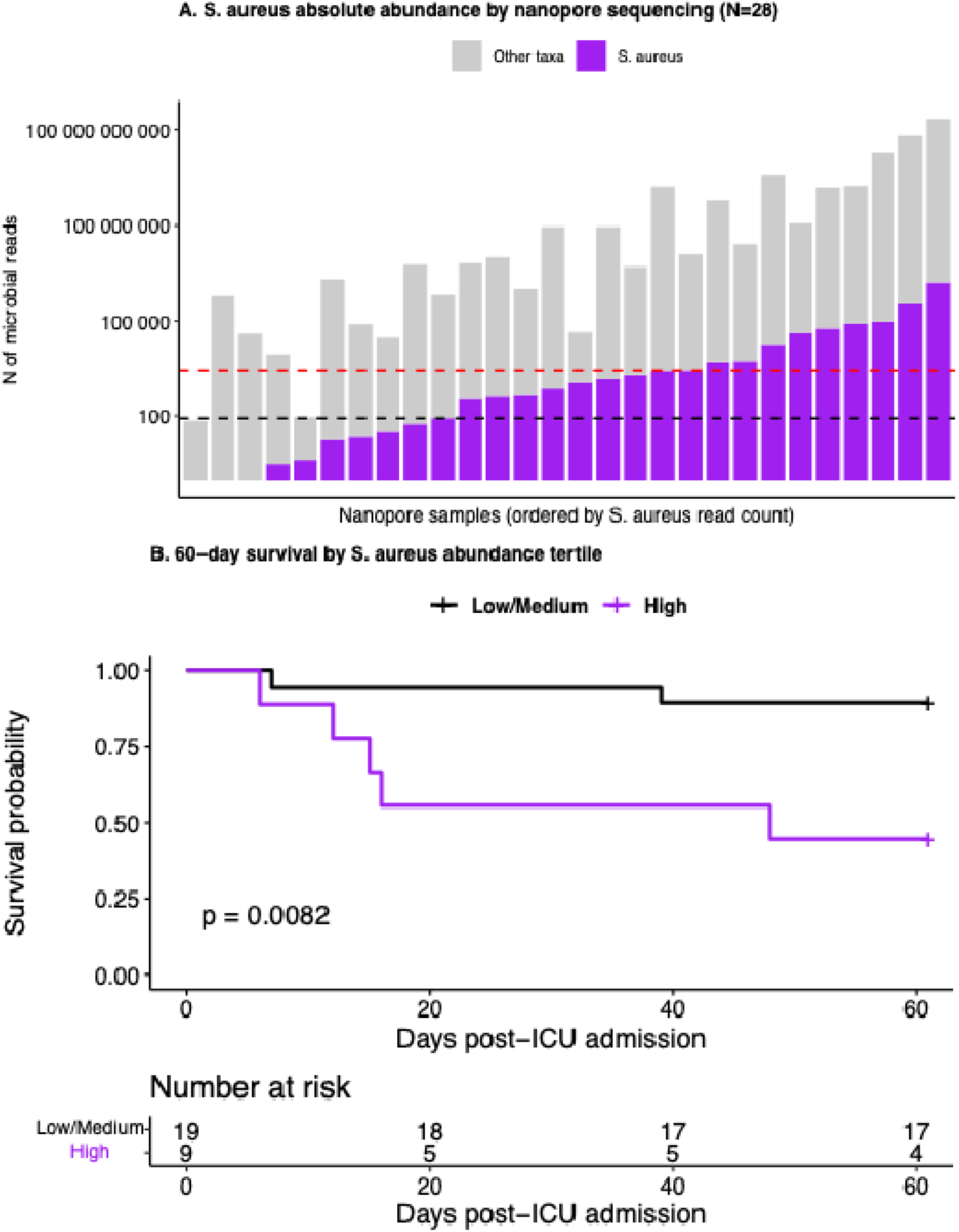
Nanopore Metagenomic Sequencing Reveals *S. aureus* Abundance Heterogeneity and Predicts Mortality. (A) Stacked bar plot showing absolute *S. aureus* read counts (purple) and other taxa (gray) from nanopore metagenomic sequencing of endotracheal aspirate samples (n=28), ordered by *S. aureus* absolute abundance (log10 scale). Black dashed line indicates the lower tertile boundary (84 reads); red dashed line indicates the upper tertile boundary (2,714 reads) defining high abundance. (B) Kaplan-Meier survival curves for 60-day mortality stratified by tertile of absolute *S. aureus* read count. Patients with high abundance (purple, n=9) demonstrated significantly worse survival than those with low/medium abundance (black, n=19), with 55.6% versus 89.5% survival at 60 days (log-rank p=0.008). Numbers at risk shown below. In multivariable Cox proportional hazards analysis adjusting for age, sex, and methicillin resistance status, high abundance remained independently associated with mortality (adjusted HR 11.23 [95% CI 2.25–55.9], p=0.003).

### Virulence Gene and Antimicrobial Resistance Profiling

Nanopore mNGS enabled culture-independent assessment of antimicrobial resistance genes and virulence factors in a subset of 19 patients with sufficient sequencing depth for functional annotation. Detection of *mecA*, the gene conferring methicillin resistance, demonstrated perfect specificity with no false-positive calls: all phenotypic MSSA isolates (13/13) were correctly classified as *mecA*-negative (Figure 5A). Sensitivity was limited, with *mecA* detected in only 2 of 6 (33.3%) phenotypic MRSA cases, which was likely constrained by limited sequencing depth.

**Figure 5.**
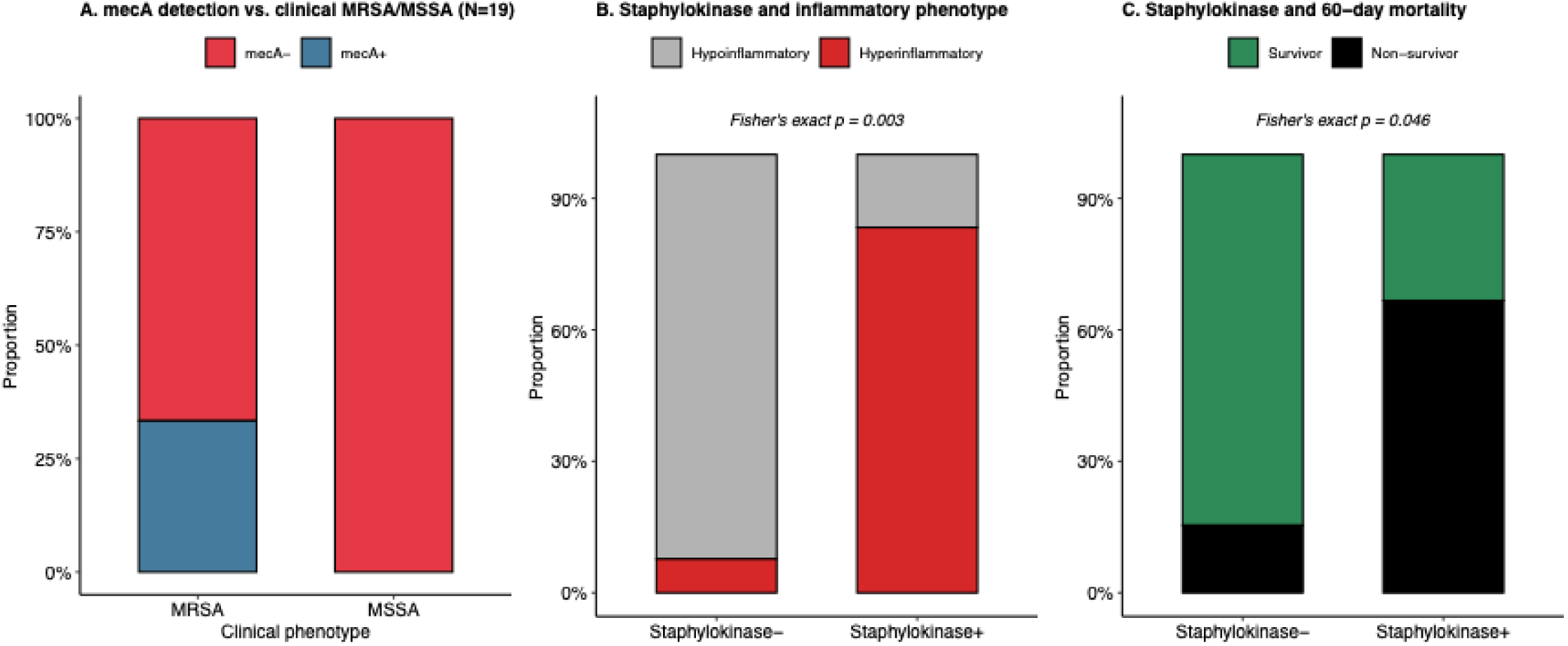
Antimicrobial Resistance and Virulence Gene Detection by Nanopore Metagenomic Sequencing. (A) Proportion of *mecA* gene detection (present/absent) stratified by clinical phenotypic classification (MRSA vs. MSSA) in 19 patients with sufficient sequencing depth for functional annotation. Nanopore sequencing demonstrated 100% specificity (0/13 false positives in MSSA cases) but limited sensitivity (2/6, 33.3%, in MRSA cases; Fisher’s exact p=0.003). (B) Proportion of hyperinflammatory versus hypoinflammatory subphenotype classification stratified by staphylokinase gene detection. Staphylokinase-positive patients were significantly more likely to be classified as hyperinflammatory (5/6 vs. 1/13; Fisher’s exact p=0.003). (C) Proportion of 60-day survival status stratified by staphylokinase gene detection. Staphylokinase-positive patients demonstrated significantly higher 60-day mortality (4/6 vs. 2/13; OR=9.28 [95% CI 0.77–181], Fisher’s exact p=0.046).

Virulence gene profiling identified staphylokinase, delta-hemolysin, and staphylococcal enterotoxin as the detected virulence factors, occurring in varying combinations across samples. We dichotomized patients by staphylokinase detection (present in 6/19, 31.6%), given its established role in immune evasion via IgG and complement cleavage and its enrichment in high-virulence *S. aureus* lineages. Staphylokinase detection was strongly associated with the hyperinflammatory subphenotype (5/6 staphylokinase-positive vs. 1/13 staphylokinase-negative classified as hyperinflammatory; Fisher’s exact p=0.003, Figure 5B) and with significantly higher 60-day mortality (4/6 vs. 2/13; OR=9.28 [95% CI 0.77–181], p=0.046, Figure 5C).

## Discussion

In this nested cohort study of mechanically ventilated patients with culture-confirmed *S. aureus* pneumonia, we demonstrate that microbiome profiling — whether by 16S rRNA gene amplicon sequencing or Nanopore mNGS — uncovers substantial and clinically meaningful microbiological heterogeneity that is masked by conventional culture methods. Despite all patients meeting the gold “aureus” standard for microbiological diagnosis of *S. aureus* pneumonia, *Staphylococcus* relative abundance varied dramatically, ranging from undetectable to near mono-detection. Critically, this heterogeneity was not attributable to random sampling variability or technical artifacts, as the same prognostic signal emerged across two distinct sequencing platforms, different time points, and independent analytical pipelines: patients with higher *Staphylococcus* abundance — whether measured by relative abundance in 16S profiles or absolute read counts in Nanopore mNGS — demonstrated significantly worse 60-day survival, even after adjusting for age, sex, and methicillin resistance.

These findings challenge the conventional interpretation of positive respiratory cultures in pneumonia. Clinicians reasonably assume that recovery of a typical respiratory pathogen like *S. aureus* from LRT cultures indicates that the organism is present in sufficient abundance to play a pathogenic role in the infection. Our results demonstrate that this assumption is frequently incorrect: despite uniform culture positivity, *S. aureus* comprised an undetectable or negligible fraction of the respiratory microbiota in many cases, while only one-third demonstrated the ecological dominance that culture growth would suggest. Our findings align with emerging evidence from VAP cohorts analyzed by complementary sequencing approaches. Two recent studies—one utilizing 16S rRNA gene sequencing in 90 patients with suspected VAP,(19) the other employing RNA metatranscriptomics in 104 VAP events(21)—independently reported the same paradoxical pattern for *Staphylococcus*: frequent culture positivity despite low relative abundance (<25%) in the majority of sequence-based microbiome profiles. While technical challenges in DNA extraction from the rigid staphylococcal cell wall have been proposed as an explanation,(19, 22) this pattern transcends cohorts, extraction methods, sequenced material (DNA versus RNA), and sequencing platforms. Instead, the convergent evidence points to a biological phenomenon: *S. aureus* demonstrates preferential growth on standard culture media,(12, 14) outcompeting fastidious or anaerobic organisms during *ex vivo* incubation and yielding results that systematically misrepresent *in vivo* microbial ecology. This phenomenon likely extends to PCR-based diagnostic panels,(23) which detect presence/absence but cannot quantify ecological context.

Critically, we demonstrate that this microbiological heterogeneity has direct prognostic implications. Patients with true *Staphylococcus* dominance experienced significantly higher mortality, even after adjusting for traditional risk factors, while culture-positive cases with minimal abundance had substantially better outcomes. Culture positivity alone thus conflates distinct biological states: high-burden infection where *S. aureus* truly dominates the respiratory ecosystem, and low-burden detection where culture amplifies a minor organism to clinical prominence.

The host inflammatory response patterns we observed provide biological plausibility for the prognostic significance of microbiota composition. Patients with *Staphylococcus* dominance demonstrated dysregulated inflammatory signaling both systemically and in the LRT. Although differences in plasma sTNFR-1 levels and hyperinflammatory subphenotype classification showed trends that did not reach statistical significance, likely due to sample size, the LRT response was markedly abnormal: ETAs from patients with dominance showed markedly elevated pentraxin-3 and depleted fractalkine. Pentraxin-3, produced locally by innate immune cells and epithelium,(24) serves as a sensitive biomarker of lung inflammation in pneumonia.(25) Conversely, given that fractalkine is produced by endothelial and epithelial cells and regulates leukocyte adhesion and chemotaxis in inflamed tissues,(26) reductions in its levels during severe pneumonia may reflect dysregulation of local immune homeostasis.(27)

Our findings situate *S. aureus* pneumonia within an evolving framework of integrated host-pathogen subphenotyping in critical illness.(28) Traditional subphenotyping schemes have operated under an implicit assumption that infectious insults are interchangeable across patients, with subphenotypic heterogeneity arising predominantly from host response differences.(28) This paradigm was recently challenged in a study of 8,280 critically ill patients, demonstrating that pathogen identity, burden, virulence, and anatomic source independently predicted sepsis subphenotype — with pathogen identity outperforming illness severity in subphenotype classification.(29) Extending this framework, we show that even within a single culture-confirmed species, quantitative microbial ecology stratifies patients into biologically and prognostically distinct groups. The association between *S. aureus* bacteremia, higher LRT abundance, and hyperinflammatory subphenotype classification mirrors the burden-barrier relationship in sepsis cohorts.(28) Our exploratory virulence gene data extend this further: higher virulence gene burden in hyperinflammatory patients suggests that pathogen fitness, not merely abundance, may influence the host response. These observations support a model in which sepsis heterogeneity reflects not only who is infected, but what is infecting them and how virulent it is — reinforcing the case for quantitative, culture-independent microbial characterization in precision medicine frameworks.

These findings raise several concrete questions for future investigation. In patients with low *Staphylococcus* abundance despite culture positivity, particularly those without a hyperinflammatory phenotype, de-escalation or earlier discontinuation of anti-staphylococcal therapy may be a reasonable target for prospective study, analogous to biomarker-guided antibiotic stewardship approaches in other pneumonia contexts. Conversely, the association between staphylokinase detection and the hyperinflammatory phenotype suggests that pathogen virulence profiling could eventually inform enrollment in host-directed immunomodulatory trials — for example, corticosteroid or anti-cytokine trials in sepsis and ARDS could be enriched for or stratified by pathogen burden and virulence characteristics rather than treating all pneumonia-associated critical illness as biologically uniform.(28)

Our nanopore mNGS results offer a preliminary, hypothesis-generating signal for culture-independent resistance profiling rather than a validated diagnostic approach at this stage. Detection of *mecA* demonstrated perfect specificity but limited sensitivity (33.3% among phenotypic MRSA cases), a finding directly attributable to low *S. aureus* sequencing depth in discordant cases. While we could not identify a precise depth threshold above which sensitivity improves given our small sample size, the association between sequencing depth and detection success suggests that optimized host-depletion protocols or deeper sequencing could meaningfully improve performance in future work.(30–32)

Several limitations warrant consideration. This single-center study enrolled a modest sample size, particularly for nanopore mNGS (n=28) and virulence gene subgroup analyses (n=19), limiting statistical power for some comparisons; nevertheless, the consistency of the core abundance-mortality relationship across two independent sequencing platforms strengthens confidence in this finding. We cannot fully exclude that comorbidity burden or immunosuppression status, which differed modestly across co-infection groups, contributed to some observed mortality differences. Whether toxin-suppressing antibiotics (e.g., linezolid, clindamycin) modify the relationship between staphylokinase detection and host response could not be examined, as only one patient in the virulence gene subset received such therapy—an important question for future, larger studies given the biological rationale for toxin suppression independent of bactericidal activity. Prospective validation of our findings and thresholds in larger, independent cohorts with formal confounder adjustment is needed before these results can inform clinical practice.

In conclusion, metagenomic profiling reveals that culture-confirmed *S. aureus* pneumonia encompasses substantial biological heterogeneity invisible to conventional diagnostics. *Staphylococcus* dominance of the LRT microbiome — reproducibly quantifiable by both 16S amplicon sequencing and Nanopore mNGS — identifies a high-risk phenotype characterized by dysregulated local and systemic inflammation and significantly worse 60-day survival. The convergence of microbial dominance, bacteremia, and virulence gene burden within the hyperinflammatory subphenotype suggests that pathogen ecology at the infection source is not epiphenomenal but mechanistically linked to clinical trajectory. As sequencing technologies mature and host-depletion protocols improve, integrating quantitative metagenomic data alongside conventional diagnostics offers a tractable path toward pathogen-informed precision medicine in critical illness.

## Acknowledgements

We wish to thank Ms. Melissa Saul and Dr. Christopher Horvat for assistance with data extractions and management. We also thank the patients and their families who have enrolled in the University of Pittsburgh Acute Lung Injury Registry, as well as the physicians, nurses, respiratory therapists and other staff at UPMC Presbyterian, Shadyside and East Hospitals intensive care units for assistance with coordination of patient enrolment and collection of patient samples.

## Data Availability Statement

The 16S rRNA gene amplicon sequencing data analyzed in this study are deposited in the NCBI Sequence Read Archive under BioProject accession PRJNA595346, available at https://www.ncbi.nlm.nih.gov/bioproject/PRJNA595346. Nanopore metagenomic sequencing data are deposited under BioProject accession PRJNA554461, available at https://www.ncbi.nlm.nih.gov/bioproject/PRJNA554461. De-identified clinical metadata and derived analytic datasets, together with the analysis code used to generate the tables and figures in this manuscript, are available at [GitHub repository URL]. Raw clinical data are not publicly available due to institutional data use restrictions but may be made available by the corresponding author upon reasonable request and execution of an appropriate data use agreement.

## AI Disclosure Statement

Claude (Anthropic) was used to assist with streamlining and debugging R analysis code, and with editing and refining manuscript text for clarity and concision. All analytic decisions, statistical interpretations, and scientific conclusions were made by the authors, who reviewed and verified all AI-assisted content for accuracy prior to submission.

