## Supplement for "Microbiome Profiling Reveals Prognostic Heterogeneity in *Staphylococcus aureus* Pneumonia"

**Supplementary Table 1. Baseline Characteristics Stratified by Methicillin Resistance**

| **Variable** | **MRSA (N=45)** | **MSSA (N=63)** | **P-value** |
| --- | --- | --- | --- |
| **Demographics** |  |  |  |
| Age, years | 58.6 [49.1, 67.9] | 51.5 [37.9, 62.7] | 0.04 |
| Male sex | 28 (62.2) | 43 (68.3) | 0.66 |
| Body mass index | 28.4 [25.6, 36.7] | 31.1 [24.8, 35.9] | 0.82 |
| **Comorbid Conditions** |  |  |  |
| Diabetes mellitus | 20 (44.4) | 22 (34.9) | 0.42 |
| COPD | 11 (24.4) | 7 (11.1) | 0.12 |
| Congestive heart failure | 6 (13.3) | 4 (6.3) | 0.37 |
| Chronic kidney disease | 6 (13.3) | 4 (6.3) | 0.37 |
| Immunosuppression | 5 (11.1) | 7 (11.1) | 1.00 |
| Chronic liver disease | 1 (2.2) | 5 (7.9) | 0.39 |
| **Microbiology** |  |  |  |
| S. aureus growth level |  |  | 0.07 |
| Rare | 5 (11.1) | 2 (3.2) |  |
| Light | 10 (22.2) | 15 (23.8) |  |
| Moderate | 17 (37.8) | 15 (23.8) |  |
| Heavy | 13 (28.9) | 31 (49.2) |  |
| Co-infection pattern |  |  | 0.38 |
| S. aureus only | 12 (26.7) | 11 (17.5) |  |
| With other pathogens | 20 (44.4) | 17 (27.0) |  |
| With non-pathogens | 13 (28.9) | 35 (55.6) |  |
| **Vital Signs** |  |  |  |
| Temperature, °C | 36.9 [36.4, 37.5] | 37.1 [36.2, 37.6] | 0.98 |
| Heart rate, beats/min | 92.0 [79.0, 106.0] | 96.0 [80.0, 112.0] | 0.33 |
| Respiratory rate, breaths/min | 22.0 [17.0, 24.0] | 22.0 [18.0, 25.0] | 1.00 |
| Systolic blood pressure, mmHg | 113.0 [98.5, 129.5] | 118.0 [110.0, 131.0] | 0.26 |
| Oxygen saturation, % | 97.0 [95.0, 98.0] | 97.0 [94.0, 99.0] | 0.66 |
| **Clinical Laboratory Values** |  |  |  |
| White blood cells, ×10⁹/L | 13.3 [10.5, 17.6] | 12.7 [8.4, 16.9] | 0.19 |
| Hemoglobin, g/dL | 10.5 [9.4, 11.8] | 11.2 [9.8, 13.4] | 0.11 |
| Platelets, ×10⁹/L | 173.0 [133.0, 245.0] | 181.0 [113.0, 247.0] | 0.96 |
| Creatinine, mg/dL | 1.6 [0.8, 3.2] | 1.0 [0.7, 2.0] | 0.07 |
| BUN, mg/dL | 34.0 [19.0, 51.0] | 27.0 [15.5, 40.0] | 0.05 |
| Bicarbonate, mEq/L | 23.0 [22.0, 26.0] | 24.0 [22.0, 27.5] | 0.53 |
| **Plasma Biomarkers** |  |  |  |
| IL-6, pg/mL | 108.9 [36.7, 261.2] | 120.5 [25.5, 523.2] | 0.75 |
| IL-8, pg/mL | 18.3 [8.8, 28.1] | 19.5 [10.3, 45.3] | 0.40 |
| sTNFR-1, pg/mL | 6499.9 [3370.8, 10255.6] | 3835.9 [2374.9, 6335.2] | 0.04 |
| Pentraxin-3, pg/mL | 5789.0 [2253.7, 14990.3] | 7035.8 [2728.6, 13916.9] | 0.84 |
| **ETA Biomarkers** |  |  |  |
| IL-6, pg/mL | 4050.8 [57.4, 8297.4] | 75.7 [21.6, 378.7] | 0.17 |
| Pentraxin-3, pg/mL | 77770.5 [5477.7, 549198.7] | 2297.0 [545.5, 314473.0] | 0.56 |
| **Ventilatory Characteristics** |  |  |  |
| Plateau pressure, cm H₂O | 22.0 [17.0, 25.5] | 20.5 [16.0, 24.0] | 0.38 |
| Ventilatory ratio | 1.6 [1.4, 2.1] | 1.4 [1.3, 2.0] | 0.49 |
| Worst PaO₂/FiO₂ ratio | 208.3 [151.6, 264.5] | 212.0 [150.3, 277.1] | 0.98 |
| **Inflammatory Phenotype** |  |  |  |
| Hyperinflammatory | 13 (30.2) | 11 (19.0) | 0.28 |
| **Severity of Illness** |  |  |  |
| Vasopressor use | 39 (86.7) | 54 (85.7) | 1.00 |
| Vasopressor days | 3.0 [1.0, 6.0] | 3.5 [2.0, 6.0] | 0.62 |
| **Clinical Outcomes** |  |  |  |
| Ventilator-free days | 20.0 [0.0, 24.0] | 14.0 [0.0, 23.0] | 0.72 |
| ICU mortality | 10 (22.2) | 12 (19.0) | 0.87 |
| 60-day mortality | 13 (28.9) | 20 (31.7) | 0.92 |
| Duration mechanical ventilation, days | 7.0 [4.0, 16.0] | 10.0 [4.0, 21.2] | 0.28 |

Data are presented as median [interquartile range] for continuous variables and n (%) for categorical variables. BUN, blood urea nitrogen; COPD, chronic obstructive pulmonary disease; ETA, endotracheal aspirate; IL, interleukin; MRSA, methicillin-resistant Staphylococcus aureus; MSSA, methicillin-susceptible Staphylococcus aureus; sTNFR-1, soluble tumor necrosis factor receptor 1.

**Figure S1. Clinical Culture Characteristics Do Not Predict 60-Day Survival in Culture-Confirmed S. aureus Pneumonia.** Kaplan-Meier survival curves stratified by **(A)** co-infection pattern (S. aureus only, S. aureus with other bacterial pathogens, or S. aureus with non-pathogenic organisms), **(B)** methicillin resistance status (MRSA vs. MSSA), and **(C)** semi-quantitative culture growth level (rare, light, moderate, heavy). P-values from log-rank tests are shown. None of the clinical culture characteristics showed significant associations with 60-day survival. Numbers at risk are displayed below each panel.

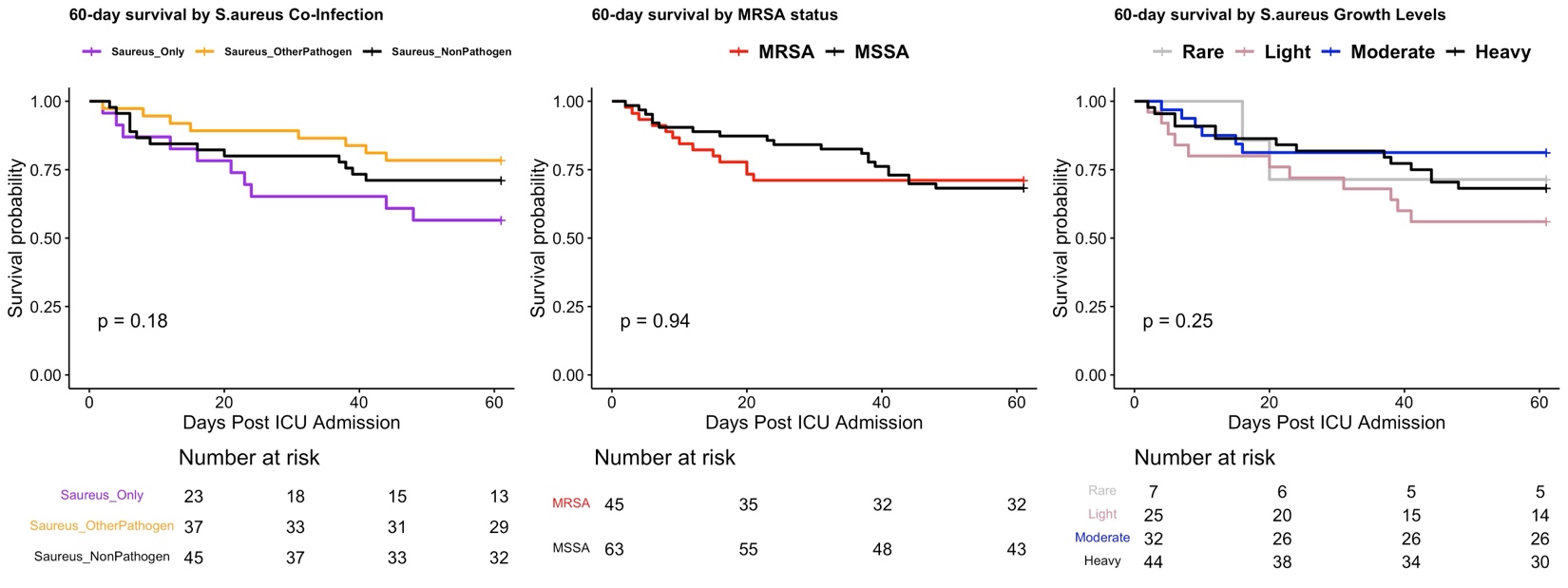

**Figure S2. Longitudinal Staphylococcus Abundance by Survival Status.**

Staphylococcus relative abundance at baseline and middle (days 3-6) timepoints in patients with serial sampling (n=19), stratified by 60-day mortality status. Survivors (green) and non-survivors (black) are shown with individual trajectories (dashed lines) and distributions (boxplots). Non-survivors demonstrated significantly higher baseline Staphylococcus abundance (p=0.01), with this difference no longer significant at the middle timepoint (p=0.11). Wilcoxon rank-sum p-values are shown for each timepoint.

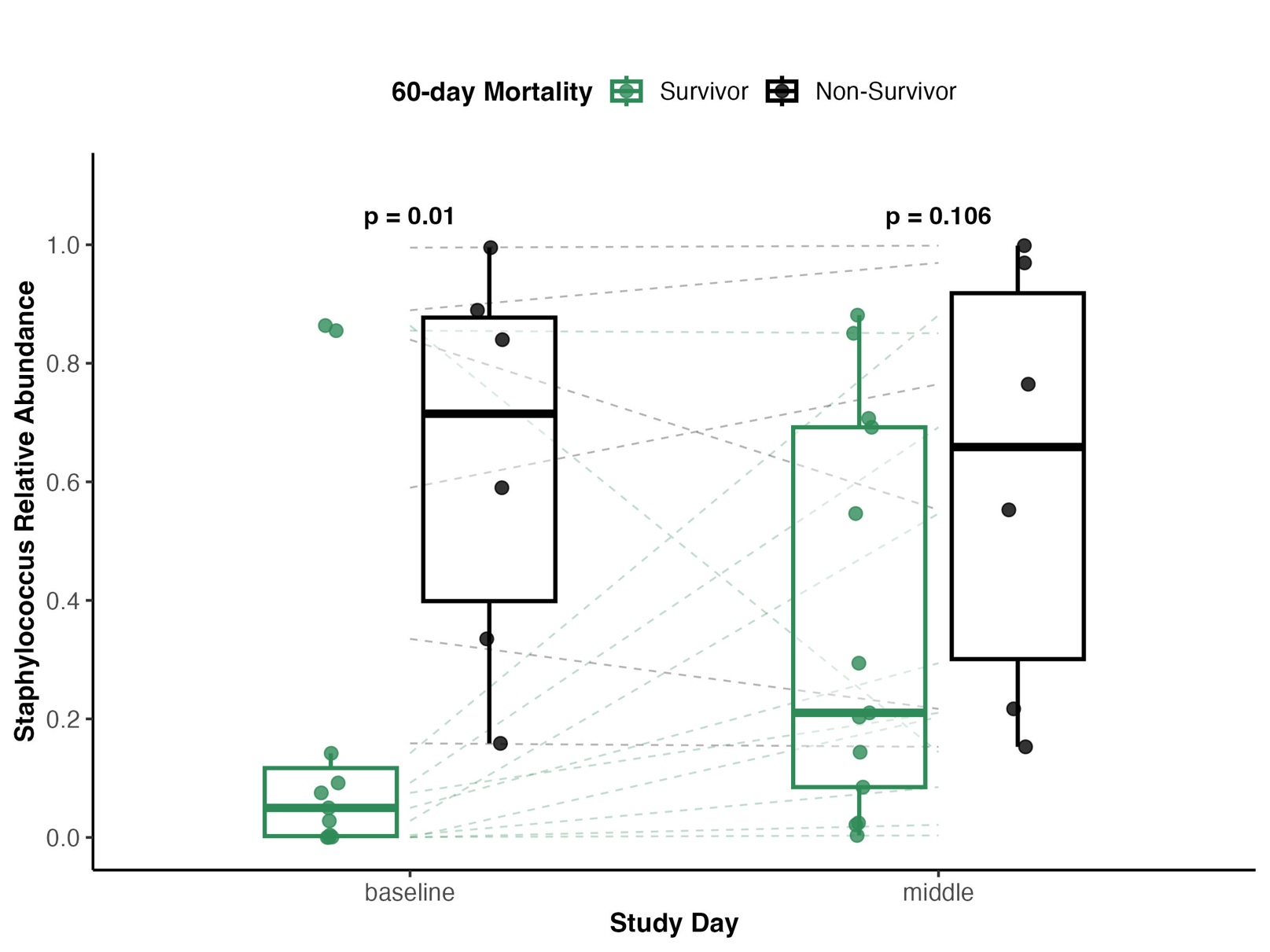

**Figure S3. Oral Staphylococcus Abundance Correlates with Lung Abundance but Does Not Predict 60-Day Survival.** (A) Scatter plot showing matched oral and lung (ETA) *Staphylococcus* relative abundance in 42 patients with baseline samples from both compartments. Oral and lung abundance were modestly but significantly correlated (Spearman ρ=0.361, p=0.019). Dashed diagonal line indicates perfect concordance; dotted lines indicate the 50% dominance threshold in each compartment. (B) Ranked lung *Staphylococcus* relative abundance (purple) with corresponding matched oral values (blue) for each participant, illustrating frequent discordance between compartments—oral swabs commonly underestimated lung *Staphylococcus* abundance in patients with high lung dominance. (C) Kaplan-Meier survival curves for 60-day mortality stratified by oral *Staphylococcus* dominance (>50% relative abundance). In contrast to the robust prognostic signal of lung dominance in the same patients (Table 2, Figure 3C), oral dominance showed no association with survival (log-rank p=0.92). Numbers at risk are shown below the curve.

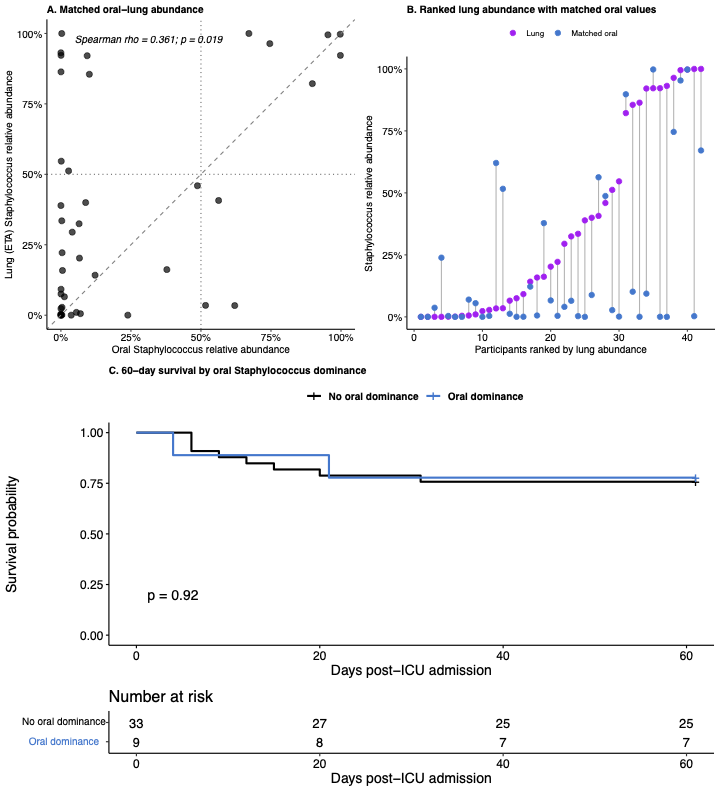

**Figure S4. Concordance Between 16S rRNA Gene Sequencing and Nanopore Metagenomic Sequencing.**

Comparison of Staphylococcus genus relative abundance by 16S rRNA gene sequencing versus S. aureus species relative abundance by nanopore metagenomic sequencing in 14 samples with both methods available. Individual samples are shown as paired barplots with SubjectID labels. Purple indicates Staphylococcus (16S genus level), pink indicates S. aureus (nanopore species level), and gray indicates other taxa. Visual inspection demonstrates strong concordance between methods, with samples showing high Staphylococcus abundance by 16S also demonstrating high S. aureus abundance by nanopore sequencing.

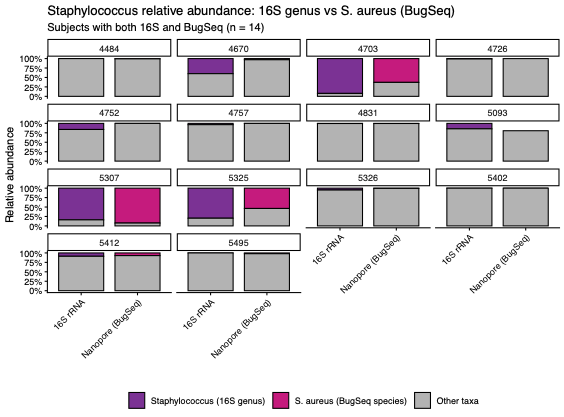

**Figure S5. Nanopore-Derived S. aureus Abundance Correlates with Clinical Features.** (A) Scatter plot showing relationship between absolute *S. aureus* read counts (x-axis, log10 scale) and *S. aureus* relative abundance among microbial reads (y-axis) from nanopore metagenomic sequencing (n=28). Strong correlation (Spearman R=0.84, p=2.7×10⁻¹⁰) indicates that abundance variation reflects true differences in bacterial burden rather than sequencing depth artifacts. Blue line shows loess smoothed fit with 95% confidence interval (gray shading). (B) *S. aureus* relative abundance by nanopore sequencing stratified by clinical culture co-infection pattern, showing no significant association at this sample size (Kruskal-Wallis p=0.11).

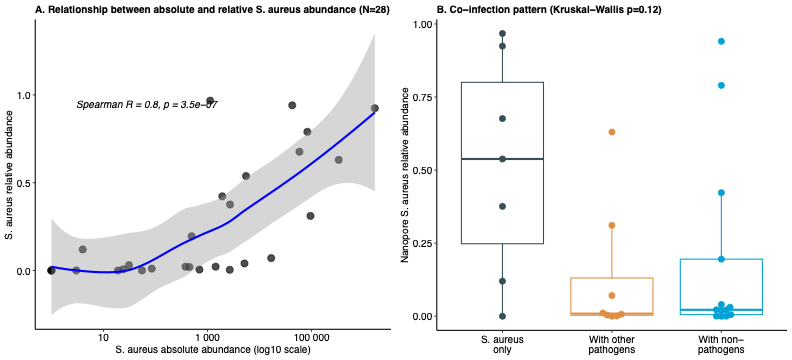
